# Microsatellites reveal high polymorphism and high potential for use in antimalarial efficacy studies in areas with different transmission intensities in mainland Tanzania

**DOI:** 10.1101/2023.12.02.23299314

**Authors:** Deus S. Ishengoma, Celine I. Mandara, Rashid A. Madebe, Marian Warsame, Billy Ngasala, Abdunoor M. Kabanywanyi, Muhidin K. Mahende, Erasmus Kamugisha, Reginald A. Kavishe, Florida Muro, Renata Mandike, Sigsbert Mkude, Frank Chacky, Ritha Njau, Troy Martin, Ally Mohamed, Jeffrey A. Bailey, Abebe A. Fola

## Abstract

**Background:** Tanzania is currently implementing therapeutic efficacy studies (TES) in areas of varying malaria transmission intensities as per the World Health Organization’s (WHO) recommendations. In TES, distinguishing reinfection from recrudescence is critical for the determination of antimalarial efficacy. Recently, the WHO recommended genotyping polymorphic coding genes (*msp1, msp2*, and *glurp*) and highly polymorphic neutral microsatellites in *Plasmodium falciparum* to adjust the efficacy of antimalarials in TES. This study assessed the polymorphisms of six neutral microsatellite markers and their potential use in TES, which are routinely performed in Tanzania.

**Methods:** *P. falciparum* samples were obtained from four TES sentinel sites, Kibaha (Pwani), Mkuzi (Tanga), Mlimba (Morogoro), and Ujiji (Kigoma), between April and September 2016. Parasite genomic DNA was extracted from dried blood spots on filter papers using commercial kits. Genotyping was performed using six microsatellites (Poly-α, PfPK2, TA1, C3M69, C2M34 and 2490) by the capillary method, and the data were analyzed to determine the extent of polymorphisms and genetic diversity at the four sites.

**Results:** Overall, 83 (88.3%) of the 94 samples were successfully genotyped (with positive results for ≥50.0% of the markers), and >50.0% (range = 47.6-59.1%) were polyclonal, with the mean multiplicity of infection ranging from 1.68 to 1.88 among the four sites. There was high genetic diversity but limited variability among the four sites based on mean allelic richness (RS = 7.48, range= 7.27- 8.03, for an adjusted minimum sample size of 18 per site) and mean expected heterozygosity (*He* = 0.83, range= 0.80-0.85). Cluster analysis of haplotypes using STRUCTURE, principal component analysis, and pairwise genetic differentiation (*FST*) did not detect any population structure, and isolates clustered independently of geographic origin. Of the six markers, Poly-α was the most polymorphic, followed by C2M34, TA1 and C3M69, while 2490 was the least polymorphic.

**Conclusion:** Microsatellite genotyping revealed high polyclonality and genetic diversity but without any significant population structure. Poly-α, C2M34 and TA1 were the top polymorphic markers and could be adopted for use in TES in Tanzania.

## Background

Malaria case management is one of the main interventions for malaria control, and together with vector control tools have significantly contributed to the reduction in morbidity and mortality that was reported between 2000 and 2015 [1]. However, this strategy has been compromised by antimalaria drug resistance which led to the withdrawal of chloroquine and SP and replacing them with artemisinin combination therapy (ACT) [2]. In 2006, Tanzania introduced ACTs with artemether-lumefantrine (AL) for the treatment of uncomplicated malaria, and the drug was officially rolled out in January 2007 [3]. AL, which is a fixed-dose combination of artemether and lumefantrine, has been effectively used for the past 16 years for the treatment of malaria [4], and studies undertaken in Tanzania have shown that it has maintained high and optimal efficacy and safety with high cure rates and minimal safety concerns [5–9]. Previous reports showed that ART-R emerged in the Mekong Subregion of Southeast Asia following deployment of ACTs and was associated with delayed parasite clearance [10,11], extended survival of ring stage [12,13] and mutations in the kelch13 (*k-13*) gene [14–16]. Until 2018, mutations associated with artemisinin resistance had not been reported in Africa [4], and ACTs retained high cure rates for the treatment of uncomplicated falciparum malaria [4]. However, recent studies confirmed artemisinin partial resistance in Rwanda with mutations at codon R561H (>5%) of the *k-13* gene and day 3 positivity rates (>10%), but AL still had sufficient cure rates (>90%) [17,18]. Similarly, artemisinin partial resistance has been reported in Uganda with mutations in the *k-13* gene at codons A675V and C469Y [19], Tanzania with 561H mutations (Ishengoma D. Unpublished data) and in Eritrea for mutations at codon R622I [20]. For lumefantrine, studies conducted in Tanzania [9] and elsewhere have reported an increase in polymorphisms in the multidrug resistance 1 gene (*mdr1*), which are associated with reduced susceptibility to lumefantrine [21]. The impacts of the polymorphism in the *mdr1* gene on the performance of AL are not clearly known, and thus, sustained surveillance is needed to monitor the performance of this important ACT and allow early detection of any emergence of resistance before its efficacy is compromised.

In Tanzania, the National Malaria Control Programme (NMCP) and its partners have been collaboratively implementing therapeutic efficacy studies (TES) since 1997 [22,23]. These TES are based on the WHO standard protocol [24,25] and aim at monitoring the efficacy and safety of antimalarials used for the treatment of uncomplicated malaria in children aged six months to 10 years. For Tanzania, studies have focused on first-line antimalarial (artemether-lumefantrine – AL) and alternative artemisinin-based combination therapies (ACTs). The current alternative ACTs covered in TES include artesunate-amodiaquine (ASAQ), which is the first-line drug in Zanzibar [26], and dihydroartemisinin-piperaquine, which was included in the National Guidelines for Diagnosis and Treatment of Malaria from 2014 [22,27] According to the WHO protocol [24,28,29], TES has two components: field data and sample collection and laboratory analyses. The laboratory analyses aim at distinguishing recrudescent from new infections in patients with recurrent infections and generating data of molecular markers in the genes associated with resistance or reduced sensitivity/susceptibility of the parasites to the drugs. To distinguish recrudescent from new infection, the old WHO protocol, which was developed in 2008, recommended genotyping of three polymorphic genes, including merozoite surface protein 1 and 2 *(msp1 and msp2)* and glutamate-rich protein (*glurp*) [29]. Recently, the WHO has recommended a new protocol that recommends genotyping both *msp1* and *msp2* and one or two highly polymorphic microsatellite markers [30]. Several microsatellite makers have been utilized in studies of malaria parasites, but they differ in their level of polymorphisms and informativeness [13,31]. Of the different microsatellites, WHO recommends using poly-A and any of the other two markers, TAE1 and Pfk2. However, these markers have not been optimized in different countries to determine if they are indeed sufficiently polymorphic and sensitive and can reliably be used for genotyping within TES. This study was therefore undertaken to assess the polymorphisms of six microsatellite markers (Poly-A, PfPK2, TA1, C3M69, C2M34 and 2490) for potential use in TES in Tanzania. The findings provide important information on these markers and parasite populations in the country and will facilitate future genomic studies and their application in TES and malaria surveillance as well.

## Materials and Methods

### Study sites

The samples used for this study were obtained from clinical malaria cases sampled in a TES that was conducted during and after the long rainy season between April and September 2016 [9]. It was undertaken in four geographically and epidemiologically distinct areas of Tanzania (Kibaha – Pwani, Mkuzi – Tanga, Mlimba – Morogoro and Ujiji - Kigoma), and these sites have been NMCP sentinel sites for monitoring of antimalarial efficacy since 1997 (Figure 1) [22,23]. The study sites were selected to represent distinct geographic areas of Tanzania. In Kibaha district of the coastal region (Pwani), the study was conducted at Yombo Dispensary, which is located in an area that has transitioned from high to low malaria transmission (with prevalence by mRDTs in under-fives of <10% in 2017)[32–34]. In Tanga region, the study site was Mkuzi health center, which is located in Muheza district. Areas around Mkuzi have reported a progressive decline in malaria prevalence (in individuals aged < 20 years) from over 80% in the 1990s to <10% in 2017 [35,36]. The Ujiji health center is located in Kigoma urban district of the Kigoma region. Parasite prevalence among under-fives (by mRDTs) in Kigoma increased from 19.6% in 2007 to 38.1% in 2016, followed by a decline to 24.4% in 2017, but this was the highest prevalence in the country [32–34]. The fourth site of Mlimba health center (parasite prevalence among under-fives in 2017 was <10%) is located in Kilombero district of Morogoro region and has experienced a significant decline in malaria burden in the last two decades [37]. Additional details of the study sites were given elsewhere [9,38].

**Figure 1.**
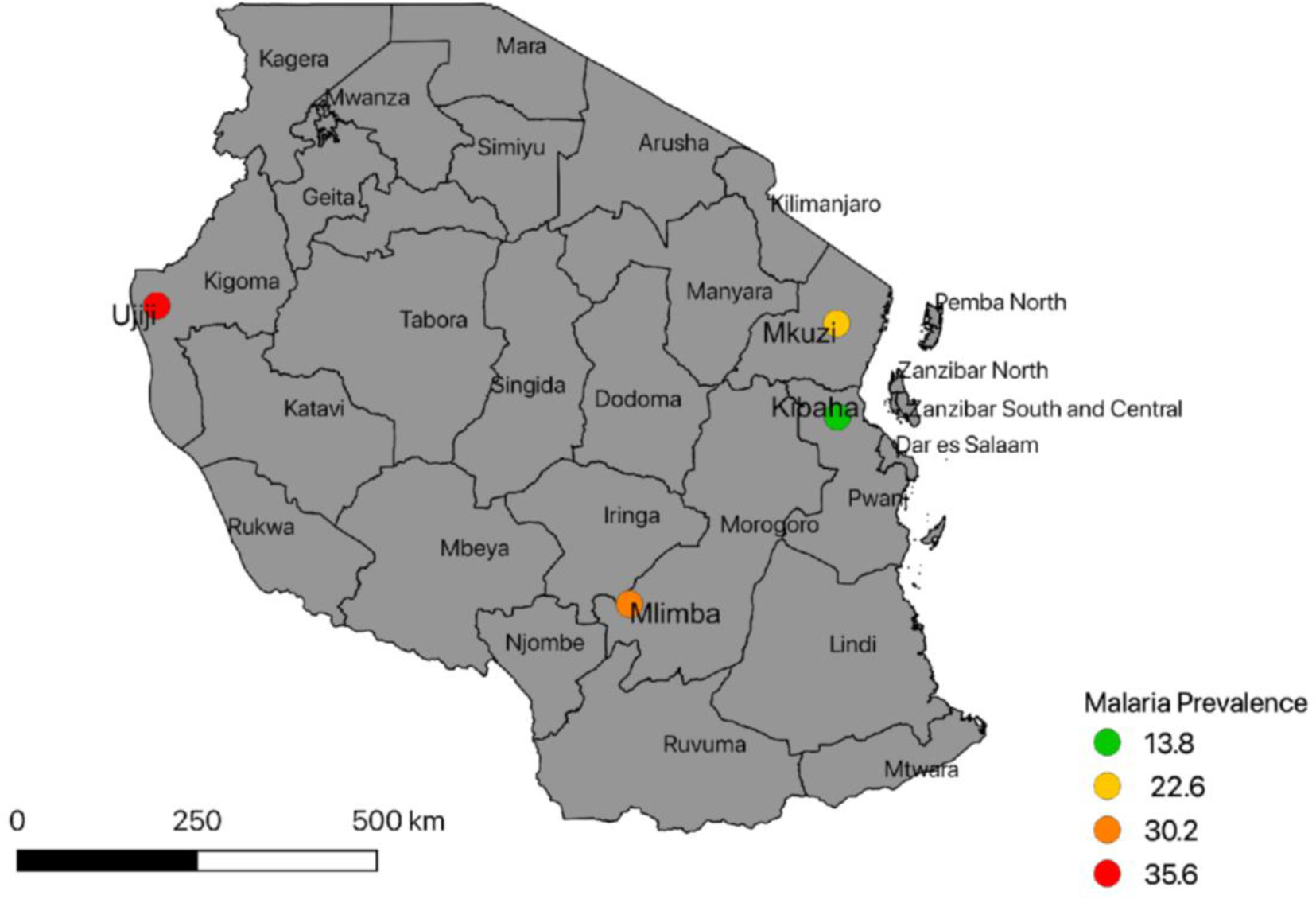
Map of Tanzania showing the four study sites of Kibaha, Mkuzi, Mlimba, and Ujiji. (Parasite prevalence data were obtained from the School Malaria Parasitological Survey of 2015[39].

### Study design and target population

Samples used for this analysis were collected during a single-arm prospective *in vivo* TES that assessed the therapeutic efficacy and safety of artemether-lumefantrine (AL) for the treatment of uncomplicated falciparum malaria and markers of artemisinin and lumefantrine resistance [9]. The study recruited 344 out of the 963 febrile children aged 6 months to 10 years who were screened according to the WHO protocol [24,25].

### Sample collection

Enrolled children were treated with AL and followed up for 28 days with clinical and parasitological assessments in the first three days post-treatment (day 1, 2 and 3) and once weekly from day 7 to 28 [9]. Thick and thin films were taken for the detection of malaria parasites during each visit. Dried blood spots (DBS) on filter papers were also collected at enrollment and from day 7 onward for molecular analyses of malaria parasites.

### Sample processing and genotyping

Parasite genomic DNA was extracted from DBS using QIAamp DNA mini-kits (Qiagen GmbH, Hilden, Germany) according to the manufacturer’s instructions. Analysis of six neutral microsatellite markers was undertaken at the Centers for Disease Control and Prevention’s (CDC) Malaria Laboratory in Atlanta, USA. A total of 94 samples were collected on day zero among patients with recurrent infections and a random selection of other samples from patients without recurrent infections collected on day zero were selected for microsatellite genotyping. These samples were analyzed to distinguish recrudescent from new infections as previously reported [9] and to determine genetic diversity in the study populations. The microsatellite markers (TA1 on chromosome 6, Poly – α on chromosome 4, PfPK2 on chromosome 12, 2490 on chromosome 10, C2M34-313 on chromosome 2 and C2M69-383 on chromosome 3) were genotyped by nested PCR for all except C2M34-313 and C2M69-383 (which were analyzed with a single step PCR). Fragment size was measured by capillary electrophoresis on ABI 3033 (Applied Biosystems) and scored using GeneMapper® Software Version 4.0 (Applied Biosystems) [40,41].

### Ethical considerations

Ethical clearance was obtained from the medical research coordinating committee (MRCC) of the National Institute for Medical Research (NIMR), while permission to conduct the study at the health facilities was sought in writing from the relevant regional and district medical authorities. Ethical clearance from the CDC was not needed because the assessments performed at the CDC Malaria Laboratory, using samples without linked identifiers (deidentified samples), were determined by the CDC Center of Global Health’s Human Research Protection Coordinator to not constitute an engagement in human subjects’ research. Informed consent (oral and written) was obtained from parents or guardians before patients were screened to assess their eligibility for possible inclusion in the study.

### Data analysis

#### Population genetic analysis

GeneScan chromatograms were analyzed using GeneMapper® Software Version 4.0 (Applied Biosystems) with an internal size standard of 350 Rox. The stutter window was set to 2.5 for 2 bp repeats, 3.5 for 3 bp repeats and 4.5 for 4 bp repeats. The stutter ratio was set to 0.4 for the four markers, and for the remaining two markers (C2M34 and C3M69), a relatively higher stutter ratio (0.6) was set, as they showed greater stuttering during manual inspection of chromatograms. A cut-off of 1,000 relative fluorescence units (RFUs) was used to distinguish true peaks from background signals. All dominant peaks (i.e., those peaks within the size range with the highest RFUs) and any additional alleles with a minimum 30% height of the dominant allele were scored. All chromatograms were inspected manually to confirm call quality. Then, samples with low RFUs were re-analysed with a minimum fluorescence of 200 RFU. Microsatellite haplotypes comprising more than 3 (50%) successfully typed markers were selected for further analysis.

For downstream population genetics analysis, multilocus microsatellite allele data were converted into different formats using CONVERT software version 1.3.1. The number of genetically distinct parasite clones (multiplicity of infection, MOI) was calculated considering the maximum number of alleles detected at any of the six microsatellite loci. The number of clones for each population was determined by summing the total number of clones per isolate. The mean MOI for each population was calculated by dividing the total number of clones detected by the number of samples. Genetic diversity was measured by calculating allelic richness (*Rs*) and expected heterozygosity (*He*) using FSTAT software version 2.9.3.2 [42]. As a measure of inbreeding within populations (non-random association of alleles), the standardized index of association (IAS) was used to measure multilocus linkage disequilibrium (LD) in each parasite population using LIAN version 3.6, applying a Monte Carlo test with 100,000 re- sampling steps [43].

STRUCTURE version 2.3.4 [44] was used to determine the number of population clusters (K) and whether haplotypes clustered according to their geographic origin. The analysis was run 20 times for K = 1 to 20, for 100,000 Monte Carlo Markov Chain (MCMC) iterations after a burn-in period of 100,000, and using the admixture model. To obtain the optimal K value, the method of Evanno *et al* [44]was used to calculate ΔK from the log probability of the data (LnP[D]) using STRUCTURE HARVESTER [45]. The STRUCTURE bar plots (ancestry coefficients) were visualized using Cluster Markov Packager Across K (CLUMPAK) [46]. Genetic differentiation between populations was measured by calculating the FST statistic according to Nei [47]. Estimation of average heterozygosity and genetic distance from a small number of individuals was performed using the pairwise.neifst function available in the hierfstat R package[48] . The Mantel test was performed to measure the associations between genetic distance and geographical distance between catchments using the Mantel function in the R adegenet package [49]. To assess haplotype relatedness, the genetic distance metric (1-pairwise allele sharing (PS)) was calculated and used to generate phylogenetic trees using the bionj Ape R package [50].

## Results

From a total of 94 *P. falciparum* samples, 83 were successfully genotyped, and all gave positive results for 3 (50%) or more microsatellite markers (Table 1). Only single infections or dominant haplotypes constructed from multiple infection data were included in downstream population genetic analyses. The number of clones per sample ranged from 1 to 4, and a total of 38 (45.8%) samples had single infections, followed by samples carrying two distinct parasite clones (n= 31, 37. 3%), three (n= 12, 14.45%), and only two samples carried four distinct clones. Overall, at least 38% of the samples in each population contained more than one parasite clone (polyclonal), and there was limited variability in the mean MOI among populations (average MOI ranged from 1.68 to 1.88) (Table 1).

**Table 1.** Population genetic metrics of four Tanzanian *P. falciparum* populations.

| Population | Prevalence (%)† | N | n | *h* | $R_s \pm SD$ | $H_e \pm SD$ | $I_A^S$ | MOI | Polyclonality (%) # |
| --- | --- | --- | --- | --- | --- | --- | --- | --- | --- |
| Kibaha | 13.8 | 25 | 21 | 19 | 8.03±2.8 | 0.85±0.12 | 0.1569** | 1.76 | 47.6 |
| Ujiji-Kigoma | 35.6 | 23 | 22 | 22 | 7.48±2.1 | 0.84±0.10 | 0.1736** | 1.88 | 59.09 |
| Mkuzi-Muheza | 22.6 | 25 | 22 | 22 | 7.79±2.4 | 0.82±0.14 | 0.1095** | 1.68 | 54.5 |
| Mlimba-Kilombero | 30.2 | 21 | 18 | 18 | 7.56±2.3 | 0.80±0.20 | 0.0903* | 1.72 | 55.5 |

*N*=total number of samples; *n*=number of samples successfully genotyped; *h*= number of unique haplotypes; $R_s$ = Allelic richness, $H_e$ = expected heterozygosity, $I_A^S$ =standard index of association as a measure of multilocus linkage disequilibrium (LD), MOI= multiplicity of infection (mean number of clones per infection); Polyclonality= proportion of samples containing more than one parasite clone †Prevalence of malaria in the study districts in the 2015 school malaria parasitological survey [10] \*=*p*-value<0.01, \*\*=*p*-value<0.001; #Polyclonality (%) refers to the proportion of infections with >1 clones.

### No significant difference in mean multiplicity of infection among populations

In malaria endemic countries such as Tanzania, individuals often carry more than one parasite clone that is genetically different, referred to as multiplicity of infections (MOI), also known as complexity of infection (COI). MOI occurs either due to repeated bites of infective mosquitoes or multiple clones in a single mosquito inoculum [51] and decreases with declining transmission. Here, we found a mean MOI of 1.73 across populations (range = 1- 4 parasite clones per sample), and there was no statistically significant difference among the four populations (Kruskal‒Wallis test) (Figure 2).

**Figure 2.**
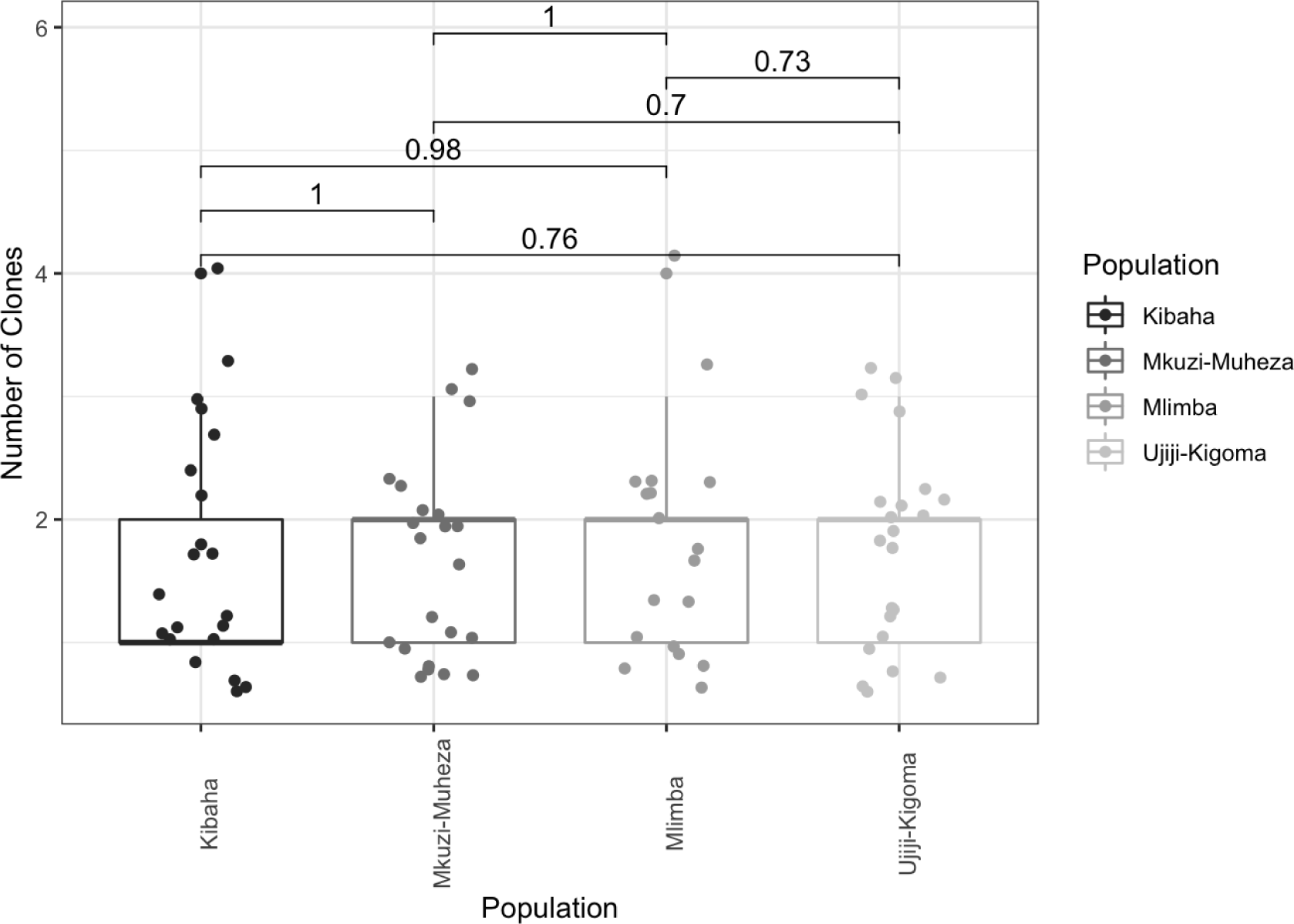
Multiplicity of infection in four Tanzanian P. falciparum **populations.** Box and whisker plots were generated from the number of clones determined for each microsatellite marker per population using R software. Dots indicate a haplotype, boxes indicate the interquartile range, the line indicates the median and the whiskers show the 95% confidence intervals. The numbers above the box plot indicate pairwise comparative p values between populations showing a lack of significant difference in MOI among the four sites.

The association between polyclonality (proportion of multiple infections) and parasite prevalence was assessed, and a positive correlation between polyclonality and malaria prevalence was observed (based on 2015 school survey data) per population (R = 0.97, *p-value* = 0.035, Spearman Rank Correlation) (Figure 3). Polyclonality was lower in Kibaha and Muheza, which had a lower prevalence compared to Ujiji, with higher polyclonality and a higher prevalence of malaria.

**Figure 3.**
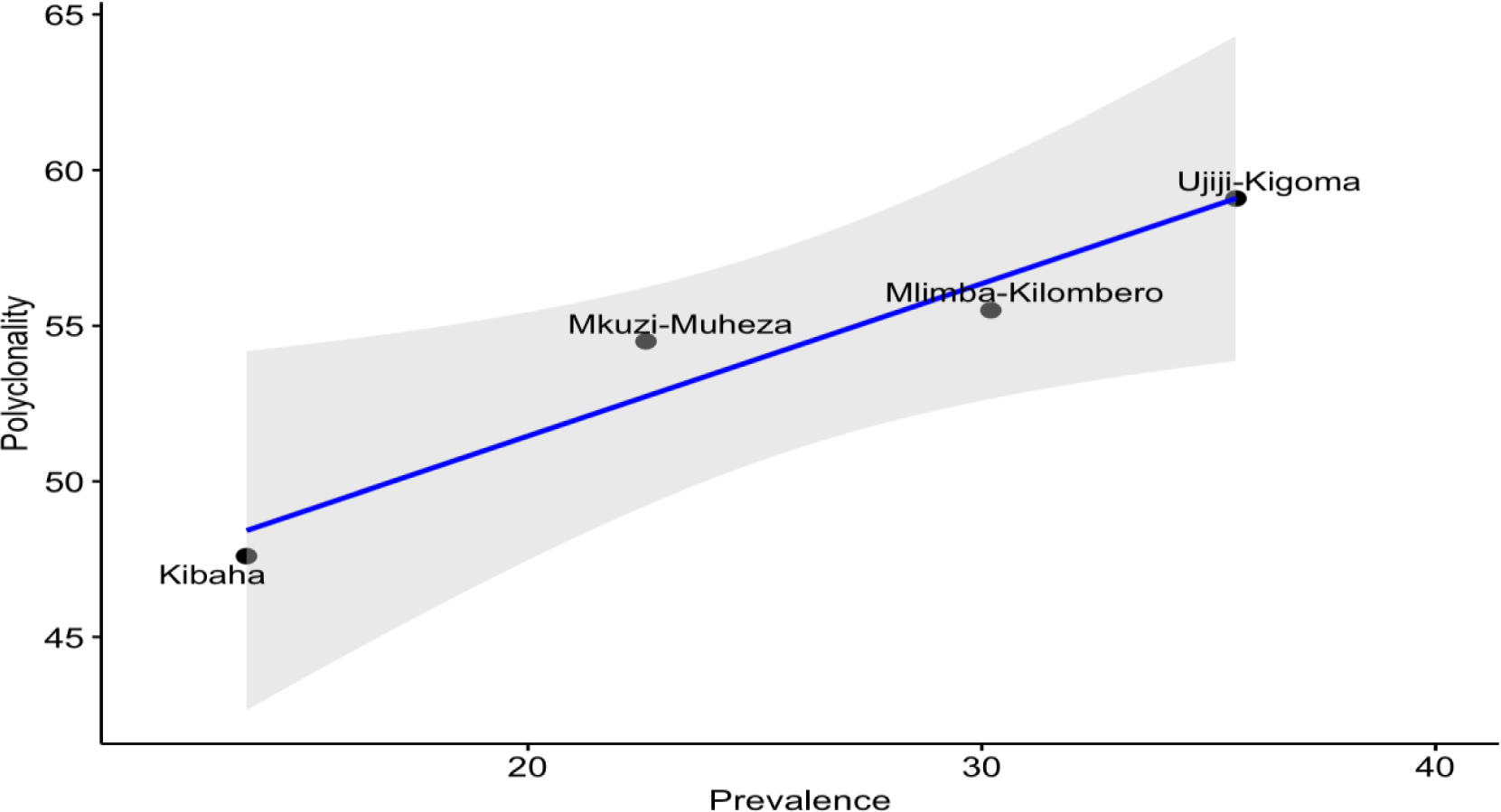
Association between the parasite prevalence (by mRDT) and proportion of multiple infections (polyclonality) in four Tanzanian *P. falciparum* populations. *The graph indicates a significant positive association between polyclonal infections and parasite prevalence across different geographic areas. Dots indicate population-level prevalence and proportion of polyclonal infections; the blue line indicates the Spearman Rank Correlation line, and the gray shaded region represents the 95% confidence interval*.

### High genetic diversity but significant multilocus linkage disequilibrium (LD)

Of the 83 multilocus haplotypes from successfully genotyped *P. falciparum* isolates, 53 (63.8%) were complete genotypes, of which 51 (96.2%) were unique and only two haplotypes were identical to each other within Kibaha population. Regardless of transmission intensity, there was high genetic diversity of *P. falciparum*, with limited variability among the four parasite populations based on allelic richness (mean RS = 7.27, range = 7.48-8.03, for an adjusted minimum sample size of 18 per site) and expected heterozygosity (mean *He* = 0.83, range = 0.80-0.85) (Table 1, Figure 4). However, according to the Index of Association (IAS) analysis, which is a measure of multilocus linkage disequilibrium (which emerges when genotypes are related), all parasite populations from the four sites showed significant multilocus LD (Table 1). This could be explained by the presence of some degree of inbreeding despite high transmission in some areas.

**Figure 4.**
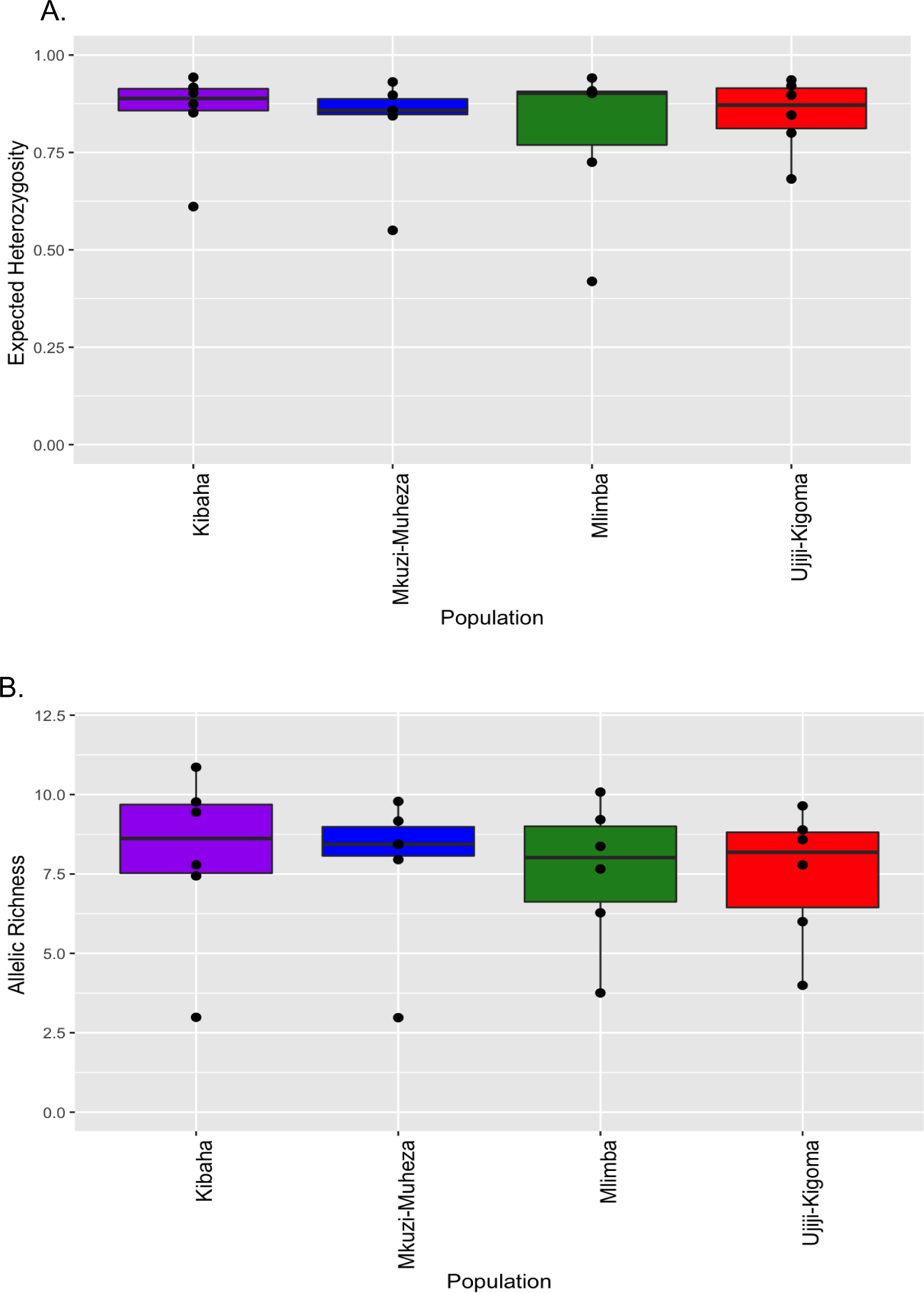
Genetic diversity (Expected Heterozygosity (A), Allelic Richness (B)) of *P. falciparum* in four geographic sites in Tanzania. *Box and Whisker plots were generated from the diversity metrics for each microsatellite marker per population using R software. Boxes indicate the interquartile range, the line indicates the median, and the whiskers show the 95% confidence intervals.*

Furthermore, the diversity of microsatellite markers was assessed, and there was high variability in the alleles present per marker (A = 3 - 13) (Figure 5), with variable frequency for the four different sites. The marker M2490 was the least diverse microsatellite, with only a 3.5 mean number of distinct alleles detected across the four populations, while PolyA was the most diverse microsatellite marker (A = 13, *He* =0.91), followed by C2M34 (A =11, *He* =0.89). These two highly polymorphic markers (polyA and C2M34) and, if needed, together with C3M69 and TA1 can be used for the detection of parasite clones in Tanzania.

**Figure 5.**
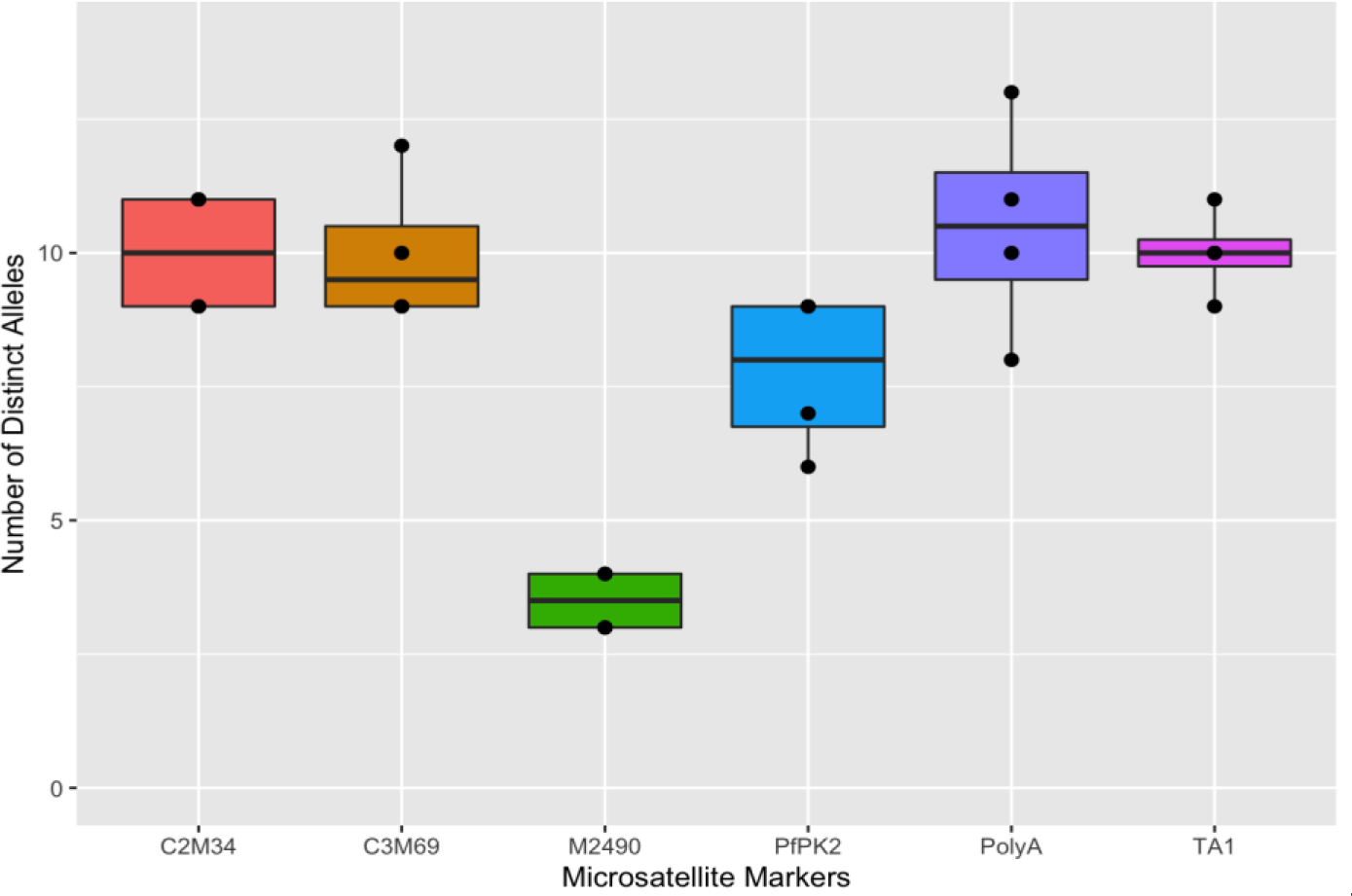
Diversity of *P. falciparum* microsatellite markers among parasites from the four sites in Tanzania. *Box and Whisker plots were generated from unique allele counts for each microsatellite marker using R software. Boxes indicate the interquartile range, the line indicates the median, and the whiskers show the 95% confidence intervals.*

### Lack of population structure and genetic differentiation

To investigate the presence of parasite population structure among the four Tanzanian sites, cluster analysis of the haplotypes was conducted using STRUCTURE version 2.3.4. No evidence of any population structure from K = 2 - 4 was detected, and the ancestry of genotypes was equally split between the genetic populations, showing no evidence of any population structure.

Further cluster analysis of haplotypes using principal component analysis (PCA, with princomp function in R package) also did not detect any signature of population structure and found no clustering of isolates by geographic origin (Figure 7).

**Figure 6.**
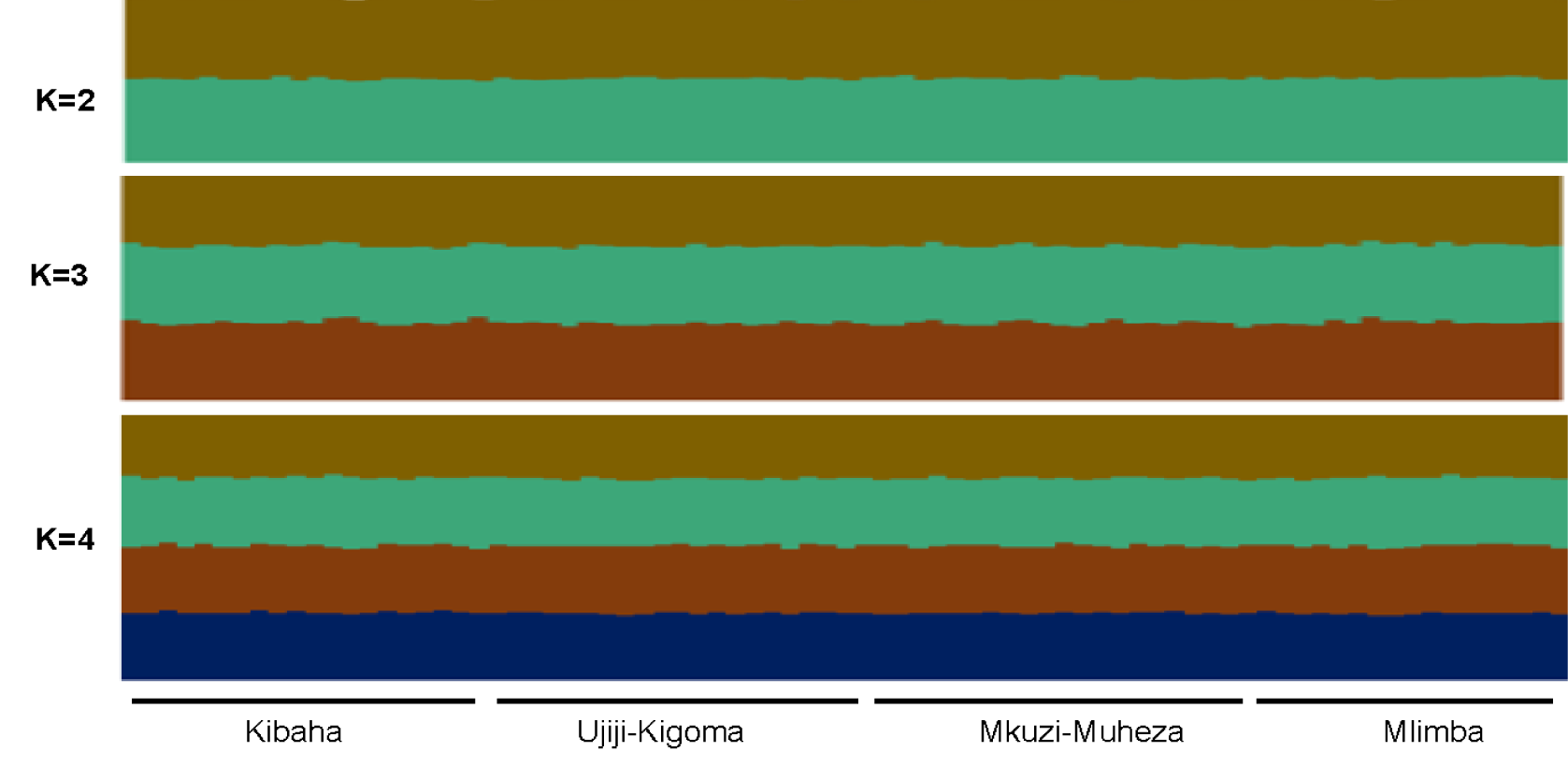
Bayesian cluster analysis of *P. falciparum* microsatellite haplotypes from the four sites of Tanzania. *Structure bar plots representing individual ancestry coefficients are shown for K=2, 3 and 4, and each vertical bar represents an individual genotype and the membership coefficient (Q) within each of the genetic populations, as defined by the different colours*.

**Figure 7.**
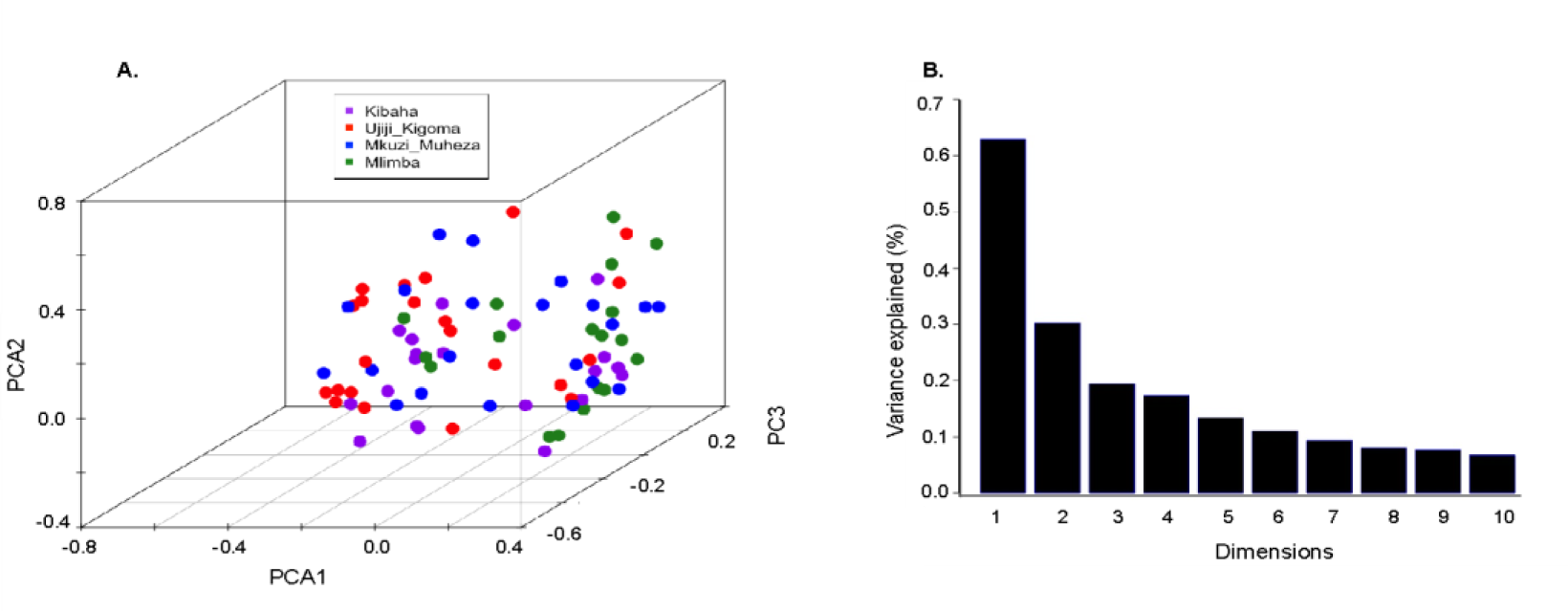
*Plasmodium falciparum* haplotype clustering. *A) Principal component analysis (PCA) of P. falciparum haplotypes. Dots indicate individual microsatellite haplotypes, and colours indicate the four sample collection sites. B) Percentage of variance explained by each principal component (PC)*.

### Gene flow and population connectivity

To assess gene flow and population connectivity, pairwise genetic differentiation based on Jost’s D metric [52] and FST according to Nei [47] were calculated among the four parasite populations using the pairwise.neifst function available in the hierfstat R package. Very low levels of genetic differentiation were observed between populations, confirming that *P. falciparum* populations from these sites are highly panmictic (Table 2). We also conducted the Mantel test to assess the correlation between pairwise genetic distance and pairwise geographic distance in km as an indication of gene flow and parasite connectivity. The differentiation of parasite populations was not statistically associated with geographical distance between populations and therefore did not fit the Isolation-by-Distance model (Mantel statistic r = 0.072, *p-value* = 0.59).

**Table 2.** Pairwise genetic differentiation among parasite populations in Tanzania.

|  | Kibaha | Ujiji-Kigoma | Mkuzi-Muheza | Mlimba |
| --- | --- | --- | --- | --- |
| Kibaha |  | 0.01 | 0.009 | 0.038 |
| Ujiji-Kigoma | 0.37 |  | 0.005 | 0.001 |
| Mkuzi-Muheza | 0.22 | 0.14 |  | 0.003 |
| Mlimba | 0.34 | 0.37 | 0.24 |  |

Top right = pairwise $F_{ST}$ , Left bottom = pairwise Jost's D

### Haplotype relatedness

To assess relatedness in *P. falciparum*, pairwise comparisons among all isolates were conducted using the dist.gene command in the R Ape package. The results showed that the majority of isolates had only one identical allele among all six markers on average, and only a few isolates shared more than 50% of the alleles (three or more alleles). Phylogenetic analysis using neighbour joining trees also confirmed a lack of population structure and geographic clustering of genotypes. However, there were more haplotypes clustering together within populations than between populations (Figure 8).

**Figure 8.**
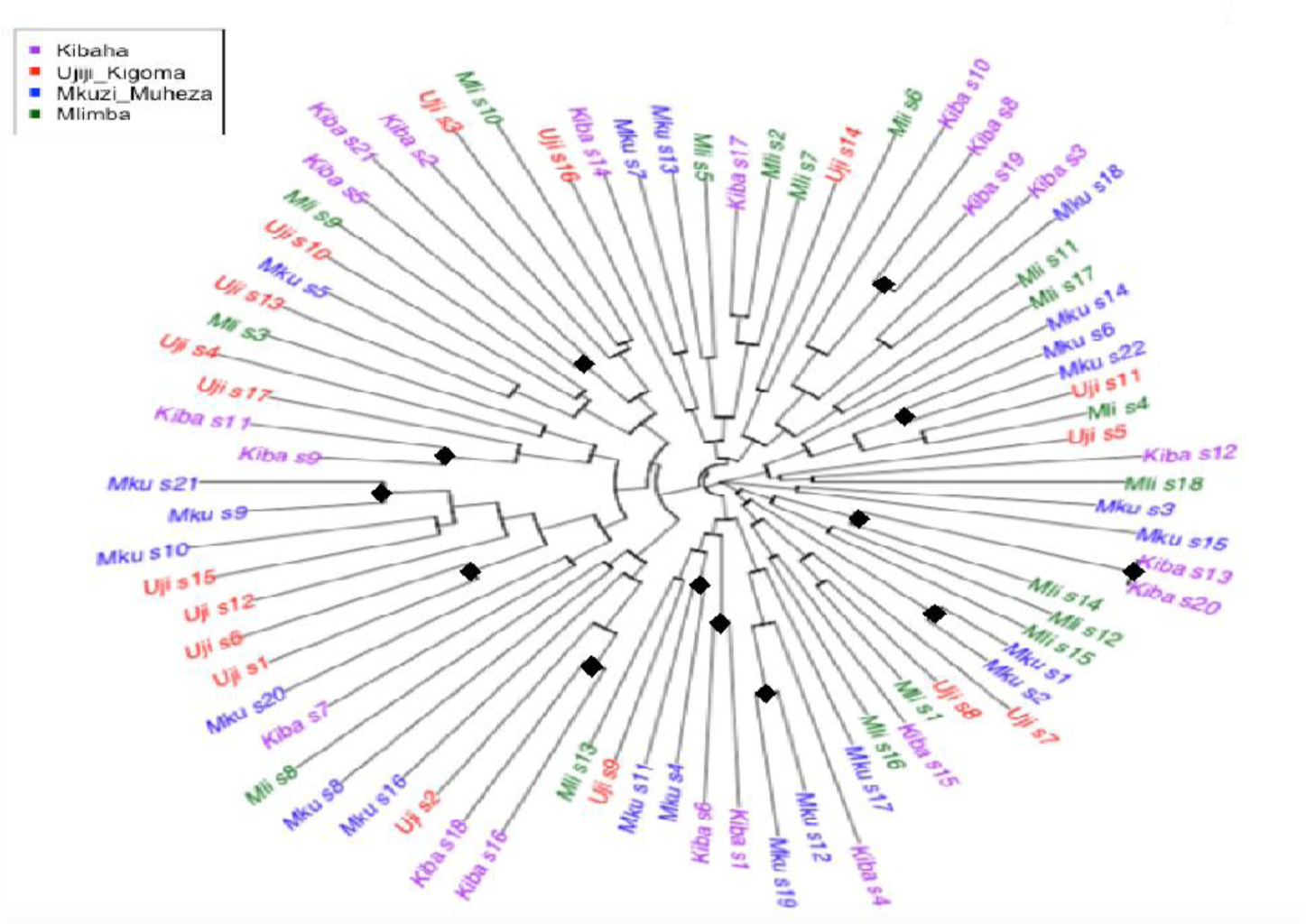
Relatedness of P. falciparum haplotypes in Tanzania. Neighbour-Joining tree showing low levels of similarity of the multilocus P. falciparum haplotypes between most isolates with similar haplotypes within populations. Tips of the NJ tree are colour coded according to the four geographic sites, and black diamonds indicate bootstrap values >50.

## Discussion

This study included samples from four geographically distinct parasite populations (located 296 to 1211 km apart) from areas with different transmission intensities to assess polymorphisms of six neutral microsatellite markers (Poly-α, PfPK2, TA1, C3M69, C2M34 and 2490) for potential use in Tanzania. It also aimed at capturing the spatial genetic diversity and population structure of Tanzanian *P. falciparum*. The findings showed that four markers (Poly-α, C2M34, C3M69 and TA1) had high diversity and could be adopted as validated markers for use in TES in Tanzania. As recently recommended by the WHO [53] and a previous study that showed that a combination of four microsatellite markers with sufficient diversity are needed in TES [54,55], these microsatellite markers can be included in the revised workflow for TES in Tanzania. The new panel should replace the old system based on genotyping of *msp1, msp2* and *glurp* for distinguishing recrudescent from new infections in ongoing TES in Tanzania. However, the areas around TES sites have increasingly reported a decline in malaria transmission in the past two decades, which suggests that continuous assessment of these and possibly other microsatellite markers will be critical. This will ensure that high-resolution markers are used and that the efficacy of antimalarials is not underestimated due to the limited discrimination power of the markers. Additional methods such as targeted amplicon sequencing can also be explored based on the capacity of the laboratory, as recently recommended [56].

This study also showed high diversity and a lack of population structure and a high level of polyclonality despite varying malaria prevalence among the study sites. The results suggest that these areas still have high malaria transmission with little evidence of a control impact on transmission dynamics. However, a significant correlation between parasite prevalence and polyclonality (as a proxy of malaria measure of transmission intensity) was detected as expected, given that in areas with higher malaria prevalence, humans are exposed to multiple mosquito bites (superinfection) or infections with multiple clones (co-transmission) [57,58]. A strong correlation between parasite prevalence and polyclonality has been reported in other studies [59–61] and needs to be monitored as a surrogate measure of potential changes in malaria transmission due to the impacts of interventions. In Papua New Guinea, *P. falciparum* MOI was associated with parasite prevalence, but the diversity of the size polymorphic markers remained high despite wide variation in prevalence at different sites [59–61]. In contrast to these findings, a study in Indonesia [62] reported lower genetic diversity, which was consistent with the low level of malaria transmission at the study sites and could be a result of longer-term sustained low transmission in this area compared to PNG and Tanzania.

Microsatellites are highly polymorphic, rapidly evolving and therefore may need long-term sustained low transmission to detect a signal of low diversity [31]. In PNG, studies have followed *P. falciparum* populations in declining transmission for over nine years and reported very minor changes in microsatellite diversity [63]. Moreover, high transmission intensity, high polyclonality and therefore high rates of recombination between distinct clones (outcrossing) might obscure the expected association between the MOI and transmission intensity (prevalence) in different transmission zones. However, in low transmission areas such as South America, studies conducted in Ecuador and Peru reported infections containing clonal parasites with clear population structure [64,65].

In addition to the MOI as a proxy for transmission intensity, estimating the extent of parasite genetic diversity and population structure is essential for a deeper understanding of malaria epidemiology and transmission dynamics as well as evaluating the impact of malaria control interventions [66]. Polymorphic markers can also be used for the detection of different parasite clones in different studies, including TES. In this study, it was shown that parasite genetic diversity was high in the four Tanzanian sites regardless of the prevalence of infection and that the respective parasite populations appeared to be highly mixed with no clear genetic structure according to geographic origin. Thus, high polymorphism at all sites and with all markers suggest that these markers (especially the three topmost) can sufficiently be used in TES to distinguish recurdescent from new infections, as recommended by the WHO [67].

Unlike the expectation that geographical isolation causes limited migration among subpopulations and geographical population structure, there was no significant genetic differentiation (measured by FST) between distant and nearby parasite populations. These results suggest high malaria transmission intensity and/or extensive parasite migration throughout the country despite significant improvement in malaria control strategies and drastic declines in malaria transmission and disease burden in recent years. These findings support previous observations where genetic diversity, geographic clustering and inbreeding with strong LD as population genetic signals are expected in low transmission areas, whereas high proportions of polyclonal infections, high diversity and panmictic parasite populations were expected in areas with high levels of transmission [31]. Generally, the levels of allelic diversity, parasite outcrossing, and gene flow are high in African populations, low in South American populations, and intermediate in Southeast Asian populations [31]. The results of this study support a situation of continuing highly endemic transmission dynamics in the country despite the expected substantial impact of recent interventions on parasite prevalence in Tanzania.

The observed differences between this and recent studies, which were conducted in Tanzania and showed population structure among parasites from different parts of the country[68,69], could be due to the markers used, SNPs and WGS data compared to microsatellites used in the current study. Validation of microsatellite markers for surveillance is important because they have been the gold standard tool for malaria parasite population genetics for many years. Furthermore, they are cheaper and easier to access for resource-limited laboratories. Ongoing and future studies will test different markers to increase the resolution and robustness of capturing different population genetics metrics that will be useful in assessing the impact of interventions and progress toward malaria elimination in Tanzania. Additionally, optimization of markers for molecular genotyping of samples collected in TES needs to be pursued as recently recommended [56].

In contrast to the above findings, there was significant multilocus linkage disequilibrium (LD) within populations, suggesting some level of inbreeding of related parasites and repeated haplotypes, suggesting the occurrence of some clonal transmission (monoclonal infections transmitted by the mosquito vector in which parasite sexual recombination occurred between genetically identical clones, albeit within the limitations of the markers used). An additional explanation for this finding could be the presence of subpopulations within populations (Wahlund effect) [70], as the samples for this study were obtained from clinical sites where patients usually come from different geographic areas to seek medical care. Other studies in different malaria endemic countries found similar results, significant LD despite high genetic diversity and a high proportion of polyclonal infection in *P. falciparum* [71,72] and *P. vivax* [73,74]. Detection of significant LD has important implications that could facilitate inbreeding and dispersal of multilocus drug resistance haplotypes or other virulent strains. As transmission decreases in Tanzania due to intensive control activities, as shown elsewhere [63], the presence of LD combined with a lack of geographic population structure is highly likely to facilitate such events and could be a future challenge in achieving malaria elimination.

There was high diversity in each of the microsatellite markers, indicating that few highly polymorphic markers (C2M34 and PolyA) can be used to track the MOI of *P. falciparum* in Tanzania. However, the genotyped markers may have limited the resolution of the population structure. The microsatellite panel used had few markers (only six, with less than one per chromosome), many were highly polymorphic (many alleles) and they are prone to technical artefacts [75]. In addition, the sample size per population was relatively limited, with approximately 20 samples successfully genotyped per site. Therefore, some subtle differences between populations may not be detected. For example, in Kibaha, the same haplotype was found in two samples, while in all other populations, all haplotypes were unique. If more samples had been genotyped, additional repeated haplotypes may be found, and diversity measures altered somewhat. Further analysis of large numbers of samples (n>50) from additional sites (again with varying transmission intensity) and utilizing larger numbers of highly polymorphic microsatellite markers [31,76,77] and/or comparing them with SNP barcodes [78], amplicons [79–82] and WGS [83] will be needed in an effort to optimize these markers. This should also be part of the ongoing initiatives to establish a molecular surveillance platform to support policy and decision-making by the Tanzanian NMCP in their strategy to eliminate malaria by 2030. Nevertheless, the data generated provide findings of useful markers for TES and parasite populations in Tanzania, showing that there are potentially large, diverse and highly intermixing parasites despite strong reductions in infection prevalence and disease burden. The findings also provide useful baseline information for future monitoring of parasite populations in response to ongoing malaria interventions.

## Conclusion

Microsatellite genotyping revealed high polyclonality and genetic diversity but without any significant population structure. Poly-α, C2M34, C3M69 and TA1 were the top polymorphic markers and could be adopted for use in TES in Tanzania. Failure to reveal any significant population structure among parasite populations could be due to high transmission or inherent limitations of small numbers of microsatellite markers and sample size. More studies covering sites with varying transmission intensity, more samples and using other genotyping markers will be needed for establishing an effective molecular surveillance system to support implementation of TES and area-specific interventions in Tanzania and monitoring the impacts of the interventions.

## Abbreviations

ACT: Artemisinin-based combination therapy
AL: Artemether-lumefantrine
CDC: Centers for Disease Control and Prevention
DBS: Dried Blood Spot
DNA: Deoxyribonucleic acid
MRCC: Medical Research Coordinating Committee
PARMA: US PMI-supported Antimalarial Resistance Monitoring in Africa Network
PMI: US President’s Malaria Initiative
NIMR: National Institute for Medical Research
NMCP: National Malaria Control Programme
PCR: Polymerase Chain Reaction
SNP: Single nucleotide polymorphism
TES: Therapeutic efficacy study
USA: United States of America
WHO: World Health Organization

## Declarations

### Ethical approval and consent to participate

This study was approved by the MRCC of NIMR, and permission to conduct the study at the four sites was obtained from the district and regional medical authorities of the respective districts. Parents/guardians of all study participants signed an informed consent before enrollment and provided permission to use the samples for studying diversity among malaria parasites.

### Availability of data

The microsatellite data set used for this study is available and has been submitted with this paper as supplemental table 1. Additional prevalence data can be obtained from the NMCP upon making an official request.

### Authors’ contributions

DSI and RAM conceived of the idea and performed genotyping of the samples at CDC. DSI, AF and AB performed the analysis of data, interpretation of results and wrote the manuscript, with additional support from CIM, MW, BN, AMK, MKM, EK, RAK, FM, RM, SM, RN, AM and JB, who also designed the main study and took part in field data collection. DSI and CIM supervised data collection and overall implementation of the field work. TM provided technical and logistic support and was involved in project management at PATH. All the authors have read and approved the final version of the manuscript.

### Disclaimer

Ritha Njau and Marian Warsame are retired staff of the World Health Organization. They alone are responsible for the views expressed in this publication, which do not necessarily represent the decisions, policy or views of the World Health Organization. Furthermore, the findings and conclusions in this paper are those of the authors and do not necessarily represent the official position of the US Centers for Disease Control and Prevention.

## Acknowledgments

The authors wish to thank parents/guardians of children for taking part in the study. They are grateful to the health facilities’ staff, stakeholders, TES partners and local health authorities for their support. Molecular analysis was undertaken at the CDC with the support of the US President’s Malaria Initiative-supported Antimalarial Resistance Monitoring in Africa (PARMA) Network. Technical and logistic support was provided by the CDC, USAID/PARMA team (Drs. Udhayakumar Venkatachalam, Eric Halsey, Meera Venkatesan, Naomi Lucchi, Eldin Talundzic and Lynn Paxton), PMI - Tanzania (Chonge Kitojo, Gorge Greer and Erik Reaves) and NIMR staff are highly appreciated. Test drugs and filter papers were provided by the WHO through its Global Malaria Programme. Thanks to PATH teams (Seattle, WA and Washington DC, USA) for financial and logistics support. This study was sponsored by NIMR as the host institution of TES coordination on behalf of the TES Taskforce. Special appreciation to Catherine Bakaria and Salehe Mandai for helping to finetune the final version of the manuscript. Permission to publish this paper will be sought and granted by the Director General of NIMR.

## Consent for publication

Not applicable.

